# Critical and non-critical connections not differently associated with either Alzheimer’s disease or vascular pathologies

**DOI:** 10.64898/2026.02.03.26345446

**Authors:** Naomi Vlegels, Bruno M. de Brito Robalo, Alberto de Luca, Wiesje M. van der Flier, Dominantly Inherited Alzheimer Network (DIAN), Randall Bateman, Tammie L. S. Benzinger, Carlos Cruchaga, Dave M. Cash, Hiroshi Mori, Igor Yakushev, Marco Duering, Sofia Finsterwalder, Benno Gesierich, Anna Kopczak, Yael D. Reijmer, Geert Jan Biessels

## Abstract

Following observations from a pilot study that, contrary to expectations, indicated that critical white matter (WM) connections were not more vulnerable to either SVD or AD pathologies than non-critical connections, we set out to systematically evaluate the relation between these pathologies and both connections types. For patients with CADASIL (n=59), Mixed pathology (n=57) and autosomal dominant AD (ADAD; n=50) we reconstructed WM networks based on diffusion tensor imaging and subsequently defined critical and non-critical connections. Associations between AD markers (CSF Aβ^42^, p-tau levels, estimated years of onset (EYO)) and SVD markers (WM hyperintensity (WMH) volume) and both connection types were tested with linear regression analyses. WMH volume showed equally strong associations to the strength of both critical and non-critical connections. Aβ-positivity, Aβ^42^ levels, p-tau levels and EYO, while less strongly related to the strength of the WM connections, did consistently show similar effect sizes for both connection types. Sensitivity analyses using different definitions of connectivity yielded similar results. SVD burden influenced WM integrity more than AD, but we found no support for critical connections being more vulnerable to these disease effects than non-critical connections.

## 1. INTRODUCTION

Alzheimer’s disease (AD) and small vessel disease (SVD) are the two leading causes of cognitive impairment and dementia (O’Brien and Thomas, 2015). In both conditions white matter (WM) connections have been found to be disrupted, which likely contributes to cognitive dysfunction (Reijmer et al., 2013; Tuladhar et al., 2016; Heinen et al., 2018). In SVD and AD, particular attention has been given to WM connections with a critical role in the brain network, as damage to these connections may have a more pronounced effect on cognition than damage to less critical connections (Reijmer *et al*., 2016; Griffa and Van den Heuvel, 2018). Connections are considered to be critical when they link a large number of brain regions, functioning as the “highways” of the brain that are key in network integration. Due to their importance for network functioning, these critical connections have higher energy demands and associated biological costs (de Haan et al., 2012; van den Heuvel et al., 2012; Griffa and Van den Heuvel, 2018). It is hypothesized that these higher energy and maintenance requirements make critical connections more vulnerable to adverse effects of disease processes, such as SVD and AD (de Haan *et al*., 2012; van den Heuvel *et al*., 2012; Griffa and Van den Heuvel, 2018). However, to date, it is not clear whether critical connections are indeed more vulnerable to SVD or AD pathology than non-critical connections, while such insights could help to better understand disease mechanisms. Some earlier studies report a vulnerability of critical connections, without assessing non-critical connections (Daianu *et al*., 2016; Tuladhar *et al*., 2017). Other studies have reported an equal or greater vulnerability of non-critical WM connections for SVD and AD pathology (Daianu et al., 2015; Sun et al., 2019; van Leijsen et al., 2019).

In an explorative study in 186 memory clinic patients with mixed pathologies, primarily SVD and AD, we previously assessed both for critical and for non-critical connections how they related to white matter hyperintensity (WMH) volume, as a marker of SVD burden, and to medial temporal lobe volume as a measure of neurodegeneration in the context of AD (‘ESO-WSO 2020 Joint Meeting Abstracts’, 2020). Contrary to our expectations, effect sizes for the observed associations were very similar for critical and non-critical connections. We considered that these null effects, as well as some of the apparent contrasting results of previous studies, might have resulted from insufficient separation of effects of SVD and AD. Moreover, differences between previous studies could also result from different ways of defining critical and non-critical WM connections. We therefore designed the current study, meant to systematically compare critical and non-critical WM connections in relation to both AD and SVD, to support the hypothesis that these connection types are indeed equally affected by both pathologies. This systematic evaluation entailed evaluation of (full overview of design can be found in Supp. Table 1):

- Three independent patient samples with pure (i.e., monogenic) AD, monogenic SVD, or with mixed AD and SVD pathology. Within each of the three samples critical and non-critical connections were related to established markers of the severity of AD and SVD pathologies and effect sizes compared between these connection types
- Two definitions of WM connection importance (critical/non-critical and hubs/feeder/peripheral connections), each tested with weightings of connection strength by either (fractional anisotropy (FA) or mean diffusivity (MD)).

**Table 1.** Sample characteristics.

|  | **CADASIL**<br>**n = 59** | **Mixed**<br>**n = 57** | **ADAD**<br>**n = 50** |
| --- | --- | --- | --- |
| Age in years | 52.8 ± 9.9 | 73.8 ± 7.5 | 38.5 ± 11.7 |
| EYO, in years | - | - | -7.9 (23.6) [-36 – 11.8] |
| Female, n(%) | 41 (69) | 20 (35) | 34 (68) |
| MMSE | 30 (1) [22 – 30] | 27 (3) [19 – 30]<sup>a</sup> | 29 (2) [10 – 30] |
| CDR, n(%) |  | <sup>a</sup> |  |
| 0 | 50 (84.7) | 1 (1.8) | 34 (68) |
| 0.5 | 8 (13.5) | 40 (70.2) | 8 (16) |
| 1 | 1 (1.7) | 15 (26.3) | 5 (10) |
| 2 | 0 (0) | 0 (0) | 3 (6) |
| BPF | 79.1 (5.4) [69.9 – 87.2] | 68.4 (5.9) [62.3 – 74.9]<sup>b</sup> | 70.9 (5) [58 – 82]<sup>a</sup> |
| WMH volume, % of | 6.3 (6.7) [0.09 – 23] | 1.8 (1.9) [0.12 – 6.2]<sup>a</sup> | 0.17 (0.14) [0.08 – 0.5] |
| CSF Aβ<sup>42</sup> | - | 621 (168) [363 – 1641]<sup>c</sup> | 485 (444) [226 – 1463]<sup>d</sup> |
| CSF t-tau | - | 477 (376.5) [140 – | 91.4 (82.5) [8 – 272]<sup>d</sup> |
| CSF p-tau | - | 62.5 (42.5) [19 – 166]<sup>c</sup> | 49.7 (55.5) [14 – 131]<sup>d</sup> |

Data are presented in mean ± standard deviation, n (%) or median (IQR) [min, max].
Abbreviations: EYO = Estimated years from symptom onset; MMSE = Mini Mental State Exam; CDR = Clinical Dementia Rating scale; BPF = Brain Parenchymal Fraction; WMH = White matter hyperintensity; CSF = Cerebral Spinal Fluid; Aβ = Amyloid beta; t-tau = Total tau; p-tau = phosphorylated tau.
<sup>a</sup> = 1 missing; <sup>b</sup> = 2 missing; <sup>c</sup> = 19 missing; <sup>d</sup> = 8 missing

## 2. MATERIALS AND METHODS

### 2.1 Study population

#### 2.1.1 Autosomal dominant SVD

As an autosomal dominant SVD sample, we included 61 patients with Cerebral Autosomal Dominant Arteriopathy with Subcortical Infarcts and Leukoencephalopathy (CADASIL), recruited from a single-centre study (VASCAMY) at the Institute of Stroke and Dementia Research, LMU Munich (Duering et al., 2013; Baykara et al., 2016). The CADASIL diagnosis was confirmed by molecular genetic testing or skin biopsy (Joutel *et al*., 1987; Peters *et al*., 2005). 1 patient was excluded due to missing clinical data and 1 patient was excluded following visual inspection after MRI processing. 59 patients were included in the analyses.

The study was approved by the local review board and all patients provided written informed consent.

#### 2.1.2 Mixed pathology

We included 58 participants from the Parelsnoer study recruited at the memory clinic of the University Medical Centre Utrecht (Aalten et al., 2014). At baseline, all participants had a clinical dementia rating (CDR) scale score ≤ 1, and a Mini Mental State Exam (MMSE) of ≥ 20. Participants did not have another clinically apparent primary aetiology than AD-related neurodegenerative disease or SVD. For this study, we included all participants from the cohort with T1-weighted and diffusion-weighted scans and available data on their amyloid status. 1 patient was excluded due to the quality of their Diffusion-tensor imaging (DTI) scan and 57 patients were included in the analyses. The study was approved by the local review board and all patients provided written informed consent.

#### 2.1.3 Autosomal dominant AD

As an autosomal dominant AD (ADAD) sample, we included 51 participants from the observational study of the Dominantly Inherited Alzheimer Network (DIAN; data freeze 14 downloaded in December 2021) (Bateman et al., 2012; Moulder et al., 2013). DIAN enrols individuals with a known Autosomal Dominant Alzheimer’s disease (ADAD) mutation (*APP*, *PSEN1* or *PSEN2*) and their non-carrier siblings. In this study, we included both pre-symptomatic and symptomatic mutation carriers. Participants were eligible for this study if they had an available T1-weighted and diffusion-weighted MRI scan and if the scans were acquired on a Siemens scanner with a standard acquisition protocol for the diffusion scan. Participants had no other known neurological or psychiatric disease. 1 patient was excluded following visual inspection of the processed MRI data and 50 patients were included in the analyses. The study was approved by the relevant review boards and all patients provided written informed consent.

### 2.2 MRI data acquisition

MRI data for all samples were acquired on 3 Tesla systems and included 3D-T1 weighted, diffusion-weighted MRI, and T2-weighted fluid-attenuated inversion recovery (FLAIR) scans. MRI data for the CADASIL sample and the Mixed sample were acquired on one scanner each. The MRI data for the ADAD sample was acquired on four scanners, but all were Siemens scanners with a standardized protocol. Each study used a standardized protocol but acquisition parameters differed across studies (see Supp. Table 2). MRI protocols have been published for the CADASIL (Duering *et al*., 2018) and Mixed sample(Heinen et al., 2018). For the ADAD sample, acquisition parameters of the T1 and FLAIR have been previously published (Caballero *et al*., 2018).

### 2.3 Small vessel disease markers

We used WMH volume as primary marker of SVD burden because it is a robust, continuous marker of the disease with a large range in both genetic and sporadic forms. In ADAD, we did not perform analyses on WMH burden in relation to connection strength, because of two reasons: (1) WMH volume is low within the age-group studied (Finsterwalder et al., 2020; Table 1) and (2) there is uncertainty regarding the origin of WMH in ADAD (Lee et al., 2016; Shirzadi et al., 2023).

We determined normalized WMH volume for the CADASIL and Mixed sample by dividing WMH volume by total brain volume. For CADASIL, WMH volume was calculated by a semi-automated procedure which has been previously described (Baykara et al., 2016). In short, 3D FLAIR images were first segmented into tissue probability maps. CSF and WMH were then separated by histogram segmentation based on the Otsu method (Otsu *et al*., 1979) and lastly, WMH segmentations were manually edited and cleaned from misclassified artefacts. For the Mixed sample, WMH volume was calculated by a semi-automated procedure (Groeneveld et al., 2018). WMH were segmented using FLAIR images by the lesion prediction algorithm (http://nbn-resolving.de/urn:nbn:de:bvb:19-203731) as implemented in the Lesion Segmentation Toolbox (v 2.0.9; (www.statistical-modelling.de/lst.htm) for SPM12. All automated segmentations were checked visually and manually corrected if needed. Manual segmentations of infarcts were used to correct WMH segmentations.

### 2.4 Alzheimer’s Disease markers

We used CSF Aβ^42^ and p-tau levels as biomarkers of AD in the Mixed and ADAD sample. These markers were not available for CADASIL.

Concentrations of CSF Aβ^42^ in ADAD were measured by plate-based enzyme-link immunosorbent assay (ELISA) methods (INNOTEST™ Aβ1-42, Innogenetics, Ghent, Belgium). Hyperphosphorylated tau (P-tau) was analysed by multiplexed Luminex-based immunoassay (INNO-BIA AlzBio3, Fujirebio). For more details see Fagan et al. 2014 (Fagan et al., 2014). To establish Aβ-status, we used a cut-off of <600 pg/ml (Dakterzada et al., 2021). Additionally, estimated years from symptom onset (EYO) was assessed in the ADAD sample. EYO was determined based on the difference between the age of the participant at the visit and the age of onset of the parent from whom the participant inherited the mutation (Bateman et al., 2012).

For the Mixed sample, CSF Aβ^42^ and p-tau were available in a subgroup (n = 38). Concentrations of CSF Aβ^42^ and p-tau were analysed using ELISA methods (Innotest beta-amyloid (1-42) and Innotest hTAU-Ag; Innogenetics, Ghent, Belgium) (de Wilde *et al*., 2017). To establish Aβ-status, we used a cut-off of <640 pg/ml for the subgroup with available CSF data (Zwan et al., 2014). For the rest of the sample (n=22), [18F] Florbetaben PET data was available (part of the ABIDE study(de Wilde *et al*., 2017)) and Aβ-status was determined through visual read(de Wilde et al., 2017). Three participants had both PET and CSF data available, for all three Aβ-status was concordant between the two methods, as was expected based on the literature where concordance between PET and CSF data is usually high (Zwan et al., 2014; Nisenbaum et al., 2023).

### 2.5 Diffusion processing and network reconstruction

After visual inspection to exclude major artefacts, raw diffusion images were pre-processed using ExploreDTI(Leemans *et al*., 2009) (v 4.8.6) and MRtrix v3.0 package (Tournier *et al*., 2019). Pre-processing included: 1) Correction of signal drift (Vos *et al*., 2017) (ExploreDTI) 2) Denoising the data (‘dwidenoise; MRtrix) 3) Removal of Gibbs ringing artefacts(Kellner *et al*., 2016) (‘mrdegibbs’; MRtrix) and lastly 4) Correction for subject motion, eddy currents and susceptibility artefacts based on registration to the T1-weighted images in ExploreDTI, including rotation of the B-matrix (Leemans and Jones, 2009; Veraart *et al*., 2013). Pre-processing was followed by computing the diffusion tensors with robust estimators(Tax *et al*., 2015) from which we determined FA and MD. Subsequently we performed whole-brain tractography using constrained spherical deconvolution (CSD) (Jeurissen et al., 2011). CSD allows for the reconstruction of more complex pathways than DTI, i.e. multiple fiber directions in one voxel, as found in regions of crossing fibers (Jeurissen *et al*., 2011). Fiber tracts were reconstructed by starting seed points uniformly throughout the data at 2 mm isotropic resolution. Streamlines were terminated when they entered a voxel with a fiber orientation distribution threshold of <0.1, or when they deflected with an angle > 45°. The Automated Anatomic Labeling (Tzourio-Mazoyer *et al*., 2002) (AAL) template was non-linearly registered to each participants 3D-T1 image (in native space) using the Computational Anatomical Toolbox (CAT) 12 toolbox (version R1073, C. Gaser, Structural Brain Mapping Group, Jena University Hospital, Jena, Germany) for SPM version 12. All registrations were visually checked for errors. The AAL template consists of 90 cortical and subcortical brain regions.

Each AAL region represents a node in the brain network. Two nodes are considered to be connected if there are at least five streamlines where the endpoints are located in these regions, resulting in a 90×90 binary connectivity matrix. The threshold of five was set to remove noise and weak connections (Sarwar, Ramamohanarao and Zalesky, 2019; Buchanan *et al*., 2020). To assess the strength of each connection, we assessed mean diffusivity (MD) and fractional anisotropy (FA). To this end each connection within the binary connectivity matrix was multiplied by the MD of that particular connection to obtain the MD-weighted connectivity matrix and multiplied by the FA to obtain the FA-weighted connectivity matrix. These steps are further explained in the following sections.

### 2.6 Critical network connections

Connections within the brain network with an important topological position in the network are considered to be critical for global network functioning.(Van Den Heuvel *et al*., 2013) Earlier literature mentions two different definitions for critical network connections: 1) connections with high edge centrality (i.e. connections that participate in a high number of communication paths) (Reijmer *et al*., 2016) and 2) connections that form a direct link between hub nodes (van den Heuvel and Sporns, 2011). In the primary analysis, critical connections were defined as connections with high edge centrality. In these analyses, critical connections and non-critical connections were defined separately for each study sample. To define the connections with highest edge centrality, we first calculated the average brain network of each study sample based on the binary connectivity matrix by including those connections that occurred in at least 2/3 of the participants (de Reus and van den Heuvel, 2013). Centrality of each edge within the average network was quantified by the edge betweenness centrality (for the exact mathematical definition see Bullmore and Sporns, 2009). The 10% connections with the highest edge betweenness centrality were considered to be central network connections (Reijmer *et al*., 2016).

In sensitivity analyses, we defined critical connections as rich club connections, i.e. connections between hub nodes. These connections were selected as follows: first betweenness centrality was calculated for each patient separately and then averaged, secondly the 10 nodes with the highest average betweenness centrality were selected as hub nodes. All connections running between hub nodes were then defined as rich club connections (van den Heuvel and Sporns, 2011). All connections running to a hub node, were defined as feeder connections and all other connections were defined as peripheral connections (van den Heuvel and Sporns, 2011).

Non-critical connections were defined as connections that were neither included as central connections nor rich club connections (de Reus and van den Heuvel, 2013).

All these subtypes were weighted by MD-values in the primary analyses and FA-values in sensitivity analyses.

### 2.7 Statistical analysis

All statistical analyses were performed in R (version 3.5.1) (R Core Team, 2018).

Our objective was to detect differential relationships of critical and non-critical connections with disease burden within each sample. This was operationalized by comparing effect sizes and regression plots of the observed associations between these connections within each of the three samples. Because all analyses assessed both critical and non-critical connections as an outcome, we used a Bonferroni correction of factor 2 (i.e. p<0.025).

Associations between the independent variables, either markers of SVD burden markers or AD pathology, and connection measures (dependent variables) were assessed with robust linear regression analyses with M-estimation (implemented in the R-package MASS) (Venables and Ripley, 2002) within each sample. The only exception was for Aβ-positivity, a dichotomous measure, which was related to connection strength with a one-way ANOVA. Variables were log transformed in case of non-normal distribution (Shapiro-Wilk test). For all analyses, values were standardized within each respective disease sample. Given that age is a strong indicator of disease burden in both CADASIL and ADAD, all primary regression analyses were not adjusted for age. We performed sensitivity analyses in which we adjusted for age and sex in all samples. Given that in the ADAD sample multiple scanners were used, we have also performed sensitivity analyses in which we adjust for scanner.

## 3. RESULTS

Characteristics of the three samples are summarized in Table 1. As expected, patients with monogenic SVD and AD were considerably younger than patients from the mixed pathology sample. WHM volume was highest in CADASIL, intermediate in the Mixed sample, and low in ADAD. AD biomarkers had a large range in both ADAD and Mixed, which was expected given that ADAD consists of both symptomatic and pre-symptomatic patients and the Mixed sample consists of patients with mixed pathology.

### 3.1 SVD burden equally relates to critical and non-critical connections

In CADASIL, WMH volume was strongly associated with MD strength of critical and non-critical connections (critical: standardized beta (B) (95% confidence interval (CI)) = 0.70 (0.53 – 0.88), *p* < 0.001; non-critical: B(CI) = 0.70 (0.53 −0.88), *p* < 0.001). In the Mixed sample, WMH volume was similarly associated to MD strength of critical and to non-critical connections (critical: B(CI) = 0.71 (0.54 – 0.88), *p* < 0.001; non-critical: B(CI) = 0.79 (0.64 − 0.93), *p* < 0.001) (Fig. 1, Supp. table 3a).

**Figure 1.**
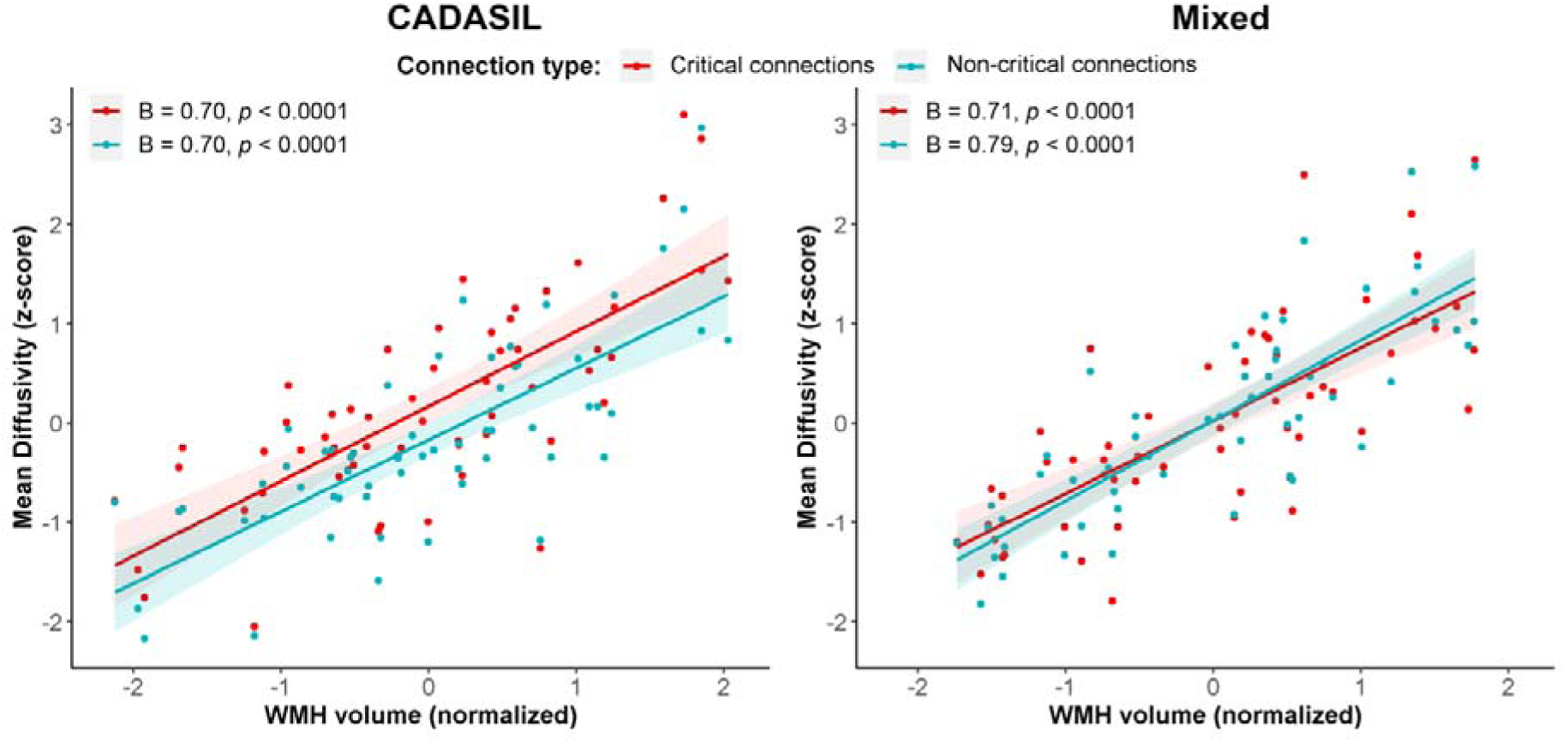
**WMH volume and MD of critical and non-critical connections in CADASIL and Mixed samples**. Shown are the associations between WMH volume and MD of critical and non-critical connections types. Shown are the standardized beta coefficients and p-values for critical and non-critical connections. WMH, white matter hyperintensity; MD, mean diffusivity.

Sensitivity analyses, which involved using FA-weighted instead of MD-weighted connections and/or using rich club connections to define the core network, as well as age and sex corrections yielded comparable findings that suggested no support of differential effects between the connection types (Supp. table 3a; Supp. table 4a).

### 3.2 AD burden not differently associated with connection types

In both ADAD and Mixed, there was no support of an association between Aβ markers and MD strength of either critical and non-critical connections. In amyloid positive ADAD patients compared to amyloid negative ADAD patients, MD was somewhat elevated, albeit not significantly, in critical (estimated marginal means (EM) (Standard Error (SE) = −0.38 (0.27), *p* = 0.17) as well as in non-critical connections (EM(SE) = −0.47 (0.24), p = 0.06) (Fig. 2b). In Mixed patients, effect sizes of amyloid positivity for the two connection types were also very similar (critical: EM(SE) = −0.25 (0.27), *p* =0.35; non-critical: EM(SE) = −0.31 (0.27), *p* = 0.25) (Fig. 2c; Supp. table 3b). In ADAD we additionally tested the association with CSF Aβ^42^ levels (critical: B (CI) = - 0.25 (−0.48 – −0.02), *p* = 0.04; non-critical: B (CI) = −0.31 (−0.54 – - 0.08), *p* = 0.01) and EYO (critical: B (CI) = 0.59 (0.37 – 0.82), *p* < 0.001; non-critical: B (CI) = 0.64 (0.42 – 0.86), *p* < 0.001). While the association between Aβ^42^ and critical connections did not survive the Bonferroni correction, effect sizes were again found to be equal between critical and non-critical connections, see Fig. 2a & 3 and Supp. tables 3c-d.

**Figure 2.**
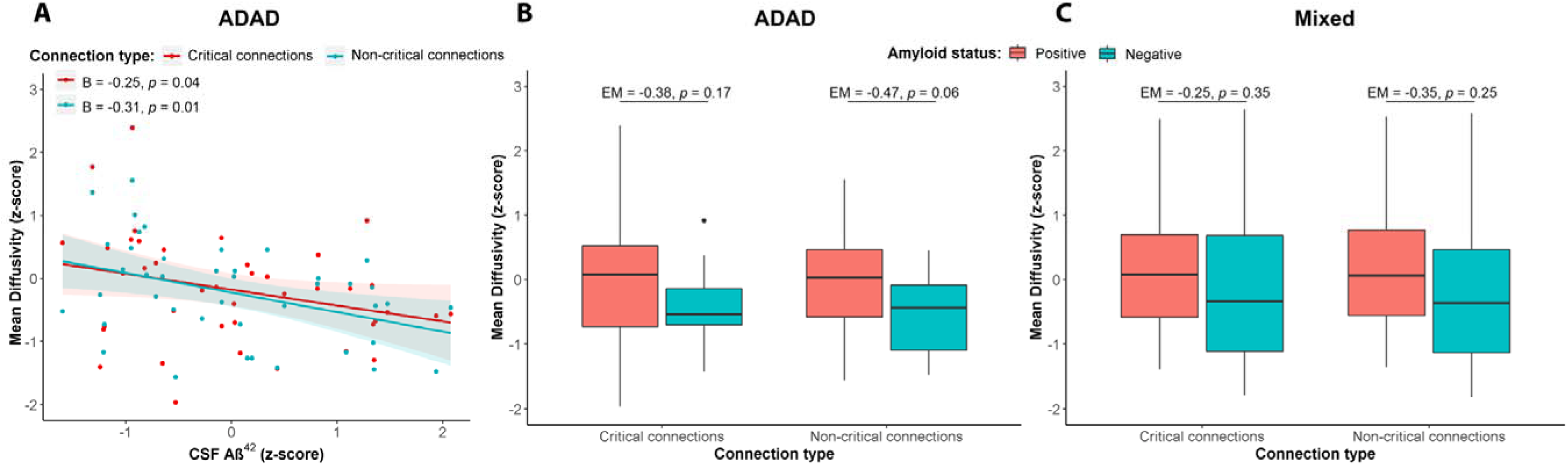
**A**β **and MD of critical and non-critical connections in ADAD and Mixed.** (A) The association between CSF Aβ^42^ levels and critical and non-critical connections. Shown are the standardized regression coefficient and p-values. (B) The association between Aβ-status in ADAD and (C) Mixed. Shown are Estimated marginal means (EM) and p-values for the difference between Aβ-positive and Aβ-negative patients. ADAD, Autosomal dominant Alzheimer’s disease; Aβ, Amyloid-beta; MD, mean diffusivity; CSF, cerebral spinal fluid; EM, Estimated marginal means.

**Figure 3.**
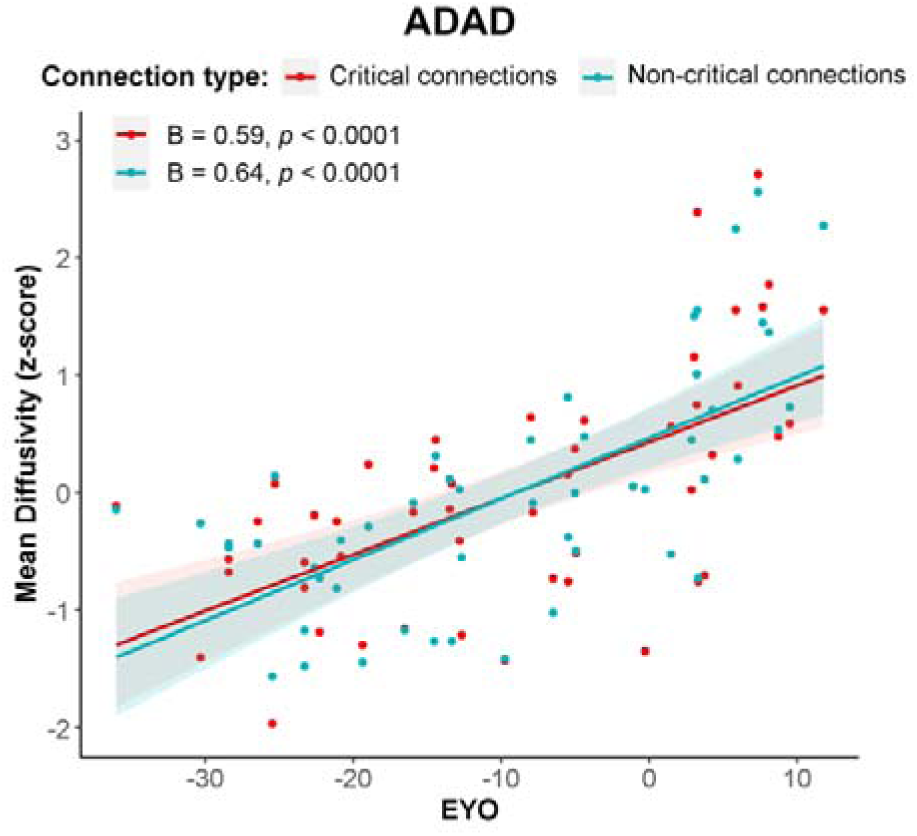
EYO and MD of critical and non-critical connections. Shown is the standardized regression coefficient and p-value for critical and non-critical connections. EYO, estimated years from symptom onset; MD, mean diffusivity.

In ADAD participants, there was a significant association between CSF p-tau and critical connections (B(CI) = 0.33 (0.13 - −0.54), *p* = 0.003) but not with non-critical connections (B(CI) = 0.21 (−0.03 – 0.45), *p* = 0.09), despite effect sizes being in the same range. In age and sex corrected analyses in the ADAD participants, CSF p-tau was not significantly related with either critical or non-critical connections, while the effect sizes did differ (critical: B = 0.23 (−0.02 – 0.48), *p* = 0.09; non-critical: B = −0.01 (−0.27 – 0.25), *p* = 0.93) (Supp Table 4e). In Mixed patients, CSF p-tau levels showed similar effect sizes in critical and non-critical connections (critical: B = 0.07 (−0.27 – 0.41), *p* = 0.7, non-critical: B=-0.07 (−0.40 – 0.26), *p* = 0.66) (Fig. 4, Supp. table 3e).

**Figure 4.**
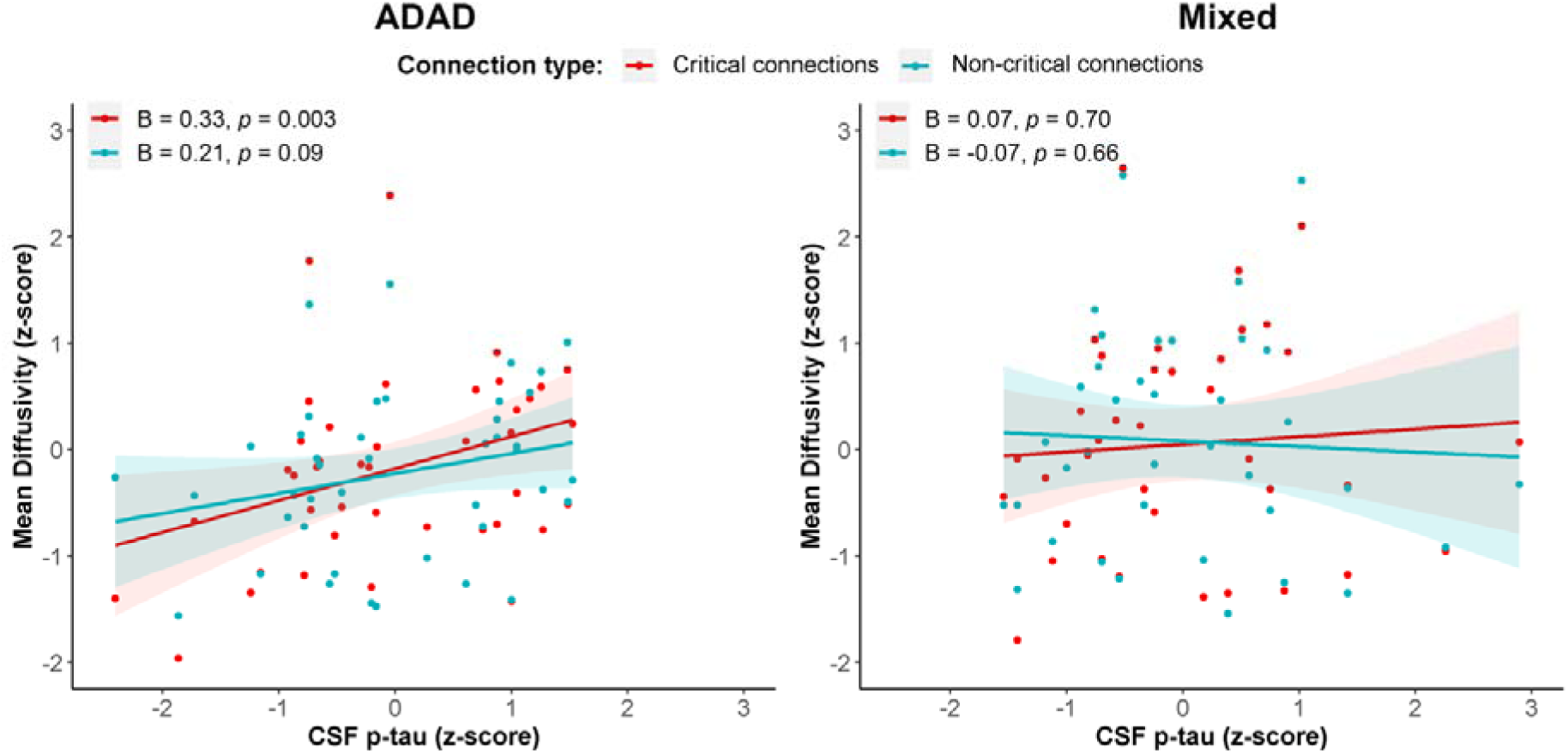
CSF p-tau levels and MD of critical and non-critical connections in ADAD and Mixed. Shown are the standardized beta coefficients and p-values is shown for critical and non-critical connections. MD, mean diffusivity; CSF, cerebral spinal fluid

Again, sensitivity analyses with FA-weighted connections and with the rich club definition of connections yielded comparable findings (Supp. tables 3b-e). The sensitivity analyses corrected for age and sex yielded, besides the CSF p-tau association, comparable findings to the main results (Supp. tables 4b-e). For the ADAD sample we have also performed a sensitivity analysis adjusting for scanner site, this resulted in similar results (Supp. table 5).

In light of the observed high concordance for the analyses of critical and non-critical connections, we also performed post-hoc analyses where we assessed the direct interrelation of the MD strength between critical and non-critical connection in each sample. This showed high inter-correlations between the connection types (all correlation coefficients > 0.84, Supp. Fig).

## 4. DISCUSSION

Using a systematic approach, including pure and mixed forms of SVD as well as AD, disease burden, based on established biomarkers, consistently related to both critical and non-critical WM connections with similar strength, supporting the hypothesis that these connection types are indeed equally affected by both pathologies.

In the different cohorts studied, due to inclusion of people at different stages of disease, we managed to obtain substantial variance in disease burden, which is an advantage for testing our hypothesis. Both in pure and mixed SVD, we found strong correlations between WMH volume, as indicator of SVD burden, and the strength of WM connections. For AD markers, we found only a weak association with strength of WM connections in both the pure and Mixed AD sample. Based on earlier literature, these differential associations between SVD and AD markers were expected (SVD: Lawrence et al., 2014; Reijmer et al., 2015; Tuladhar et al., 2016, 2017; Heinen et al., 2018); AD: Kantarci et al., 2014; Pietroboni et al., 2018; Finsterwalder et al., 2020).

In SVD, we observed a strong similarity in the relation between WMH volume and both critical and noncritical connections across our analyses. This was seen for both MD- and FA-weighted networks, as well as in sensitivity analyses with alternative definitions for critical connections. Few previous studies have specifically assessed possible differential effects of SVD on critical and non-critical connections (Reijmer *et al*., 2016; Tuladhar *et al*., 2017; Lee *et al*., 2018; van Leijsen *et al*., 2019). By comparing effect sizes for abnormalities in rich club connections in patients with SVD relative to controls, (Lee *et al*., 2018) found that patients with subcortical vascular dementia had significantly less rich club connections than controls, independent of a loss of overall connections. Using a similar analytic approach, Tuladhar et al., 2017) examined rich club organization in two cohorts of patients with SVD and found a greater loss of strength of rich club connections than that of non-rich club connections relative to controls. However, they did not perform a formal statistical test comparing the effect sizes for the two connection types. A follow-up study from the same group (van Leijsen *et al*., 2019), using a more direct approach in relating SVD burden to connectivity strength, found no differential effect of disease on critical and non-critical connections. Furthermore, in line with our findings, Reijmer et al., 2016 related WMH, lacunes and microbleeds to critical and non-critical connections in a mixed memory clinic population and also found no differential effects. These earlier findings in combination with the findings of the current systematic study make it unlikely that critical and non-critical connections are differentially influenced by SVD. One reason for this might be that SVD is a diffuse rather than a focal disease (Ter Telgte *et al*., 2018). This is also hinted towards by findings of the present study. While we found that critical and non-critical connections were almost collinear, they showed a relatively low spatial overlap, meaning that they represent different connections in the same brain areas.

For AD, we again found no indication for a differential effect on critical or non-critical connections across our analyses. As noted in the second paragraph, the association between AD biomarkers and strength of WM connections is relatively weak, compared to that of SVD burden (Finsterwalder et al., 2020). This possibly explains why the effect of AD burden on critical and non-critical connections has -to the best of our knowledge- not been studied to date. There have been few previous studies that assessed a potential differential effect on WM connections by comparing patients with controls (Daianu et al., 2015, 2016; Lee et al., 2018; Sun et al., 2019). Lee et al., 2018 examined patients with AD dementia and found a significant reduction in the number of rich club connections, independent of overall loss of connections. One study in patients with early onset AD found rich club connections to be more influenced than non-rich club connections (Daianu *et al*., 2016), while an earlier study, from the same group, which assessed patients from the ADNI cohort found no such differential effect (Daianu et al., 2015). Lastly, in patients with amnestic MCI, no differences were found in the number of rich club nodes or the strength of connections between patients and controls (Sun et al., 2019). Explanations for these differing results might be that they were all performed in slightly different groups of patients, were comparisons to healthy controls and different choices in network definitions were made. However, in our study we tested both a pure AD and a mixed pathology sample as well as four different definitions of the structural brain network and we found very similar results across all conditions. It is important to note that this study took a more direct approach by assessing the effect of disease burden, rather than absence or presence of disease, on the connections, which may also explain differences with previous studies. Yet, the results of our systematic evaluation found no differential effect of AD on critical or non-critical WM connections.

Our study has several strengths. The main strength of the current study is our systematic approach and especially the inclusion of three samples that cover the pure and mixed forms of SVD and AD. This allowed us to independently validate our findings and gave us the opportunity to study the effect of disease with minimal age-related confounders. Of note, the consistency of the findings persisted despite differences in patient characteristics and scan protocols across samples, supporting validity of the observations.

One limitation of our study is that CADASIL and ADAD are both highly selected samples, which are not generalizable to most sporadic SVD and AD populations. However, both samples gave us the unique opportunity to study SVD and AD with minimal age-related confounders. On the other hand, we also included a mixed pathology sample, which is much more heterogeneous and thus less specific but is closer to what is observed in clinical practice. Furthermore, for the CADASIL sample we had no AD biomarkers available and for the ADAD sample we did not assess SVD burden, but these were not the markers we were interested in for these groups and both SVD and AD biomarkers were available for the mixed pathology sample. Another limitation is that, while we assessed two weightings of the diffusion-based network and two definitions of critical connections, this by no means exhausts all possible options with regards to network definition. However, there is no gold standard and none of the options we assessed fundamentally changed the interpretation of the result. Interestingly, the connection types showed high inter-correlations, which could suggest that there is a high spatial overlap between these connections. We further investigated this in a representative patient, to see if this was due to a high spatial overlap between the types of connections. This did not appear to be the case; the spatial overlap was around 23%.

Finally, all three samples had a modest sample size so there might be limited power. Nonetheless, all the results that were obtained were highly consistent in terms of direction of effects and effect sizes.

Of note, while critical and non-critical WM connections appear to be equally disrupted by SVD and AD, this does not imply that both connection types are equally involved in normal or abnormal cognitive functioning. There is extensive literature, with a firm theoretical and experimental basis, that shows how network connections that have a more central role in the network are more critically involved in cognitive functioning (de Haan *et al*., 2012; Griffa and Van den Heuvel, 2018). In addition, the current observations on structural connectivity should not be equated to functional connectivity, which has a very different physiological and neuroanatomical basis. There is an extensive literature on specific functional subnetwork changes in AD, among others involving the default-mode-network in AD for example (Seeley et al., 2009).

In conclusion, we have demonstrated that critical and non-critical WM connections are similarly influenced by disease burden in SVD as well as in AD. Apparently, critical connections are not more vulnerable to these diseases, refuting earlier hypotheses.

## Data Availability

All data produced in the present work are contained in the manuscript

## ACKNOWLEDGEMENTS

We would like to thank all the researchers and the support staff from the DIAN (https://dian.wustl.edu/wp-content/uploads/2019/04/DIAN-TU-Publications_Acknowledgement_V14.pdf), Utrecht VCI study group, and CADASIL study group and study participants for their contributions to the present study. Investigators within DIAN contributed to the design and implementation of the respective studies and/or provided data but did not participate in analysis or writing of this report. A complete listing of the DIAN consortium can be found in Supplementary table 3. Members of the Utrecht VCI study group involved in the present study (in alphabetical order by department): University Medical Center Utrecht, the Netherlands, Department of Neurology: E. van den Berg, J.M.Biesbroek, M. Brundel, W. H. Bouvy, L. G. Exalto, C. J. M. Frijns, O. Groeneveld, S. M. Heringa, R. Heinen, N. Kalsbeek, L. J. Kappelle, J. H. Verwer; Department of Radiology/Image Sciences Institute: J. de Bresser, H. J. Kuijf, A. Leemans, P. R. Luijten, M. A. Viergever, K. L. Vincken, J. J. M. Zwanenburg; Department of Geriatrics: H. L. Koek; Hospital Diakonessenhuis Zeist,the Netherlands: M. Hamaker, R. Faaij, M. Pleizier, E. Vriens. Florbetaben scans of the Mixed study sample were made as part of the Dutch ABIDE project and supported by a ZonMW-Memorabel grant (project No 733050201), and through a grant of Piramal Imaging (PET scan costs). The research of GJB is supported by VICI grant 918.16.616 from NWO, the Netherlands Organization for Scientific Research.

Data collection and sharing for this project was supported by The Dominantly Inherited Alzheimer’s Network (DIAN, U19AG032438) funded by the National Institute on Aging (NIA), the German Center for Neurodegenerative Diseases (DZNE), Raul Carrea Institute for Neurological Research (FLENI), Partial support by the Research and Development Grants for Dementia from Japan Agency for Medical Research and Development, AMED, and the Korea Health Technology R&D Project through the Korea Health Industry Development Institute (KHIDI). This manuscript has been reviewed by DIAN Study investigators for scientific content and consistency of data interpretation with previous DIAN Study publications. We acknowledge the altruism of the participants and their families and contributions of the DIAN research and support staff at each of the participating sites for their contributions to this study.

## AUTHOR CONTRIBUTIONS

**NV**: Conceptualization, Methodology, Formal Analysis, Writing – Original Draft **BMdBR**: Conceptualization, Methodology, Writing – Review & Editing**, AdL**: Conceptualization, Methodology, Writing – Review & Editing**, WMvdF**: Resources, Data Curation, Funding acquisition, Writing – Review & Editing **RB**: Resources, Data Curation, Funding acquisition, Writing – Review & Editing **TLSB**: Resources, Data Curation, Writing – Review & Editing **CC**: Resources, Data Curation, Writing – Review & Editing **DMC**: Resources, Data Curation, Writing – Review & Editing **HM**: Resources, Data Curation, Writing – Review & Editing **IY**: Resources, Data Curation, Writing – Review & Editing **MD**: Resources, Data Curation, Writing – Review & Editing **SF**: Resources, Data Curation, Writing – Review & Editing **BG**: Resources, Data Curation, Writing – Review & Editing **AK**: Resources, Data Curation, Writing – Review & Editing **YDR**: Conceptualization, Methodology, Supervision, Writing – Review & Editing **GJB**: Conceptualization, Supervision, Funding Acquisition, Project administration, Writing – Review & Editing.

