## Supplemental files for "Critical and non-critical connections not differently associated with either Alzheimer’s disease or vascular pathologies"

### Supplementary Material

**Supplementary Figure** ---------------------------------------------------------------------------------------------------- **2**

#### Supplementary Figure

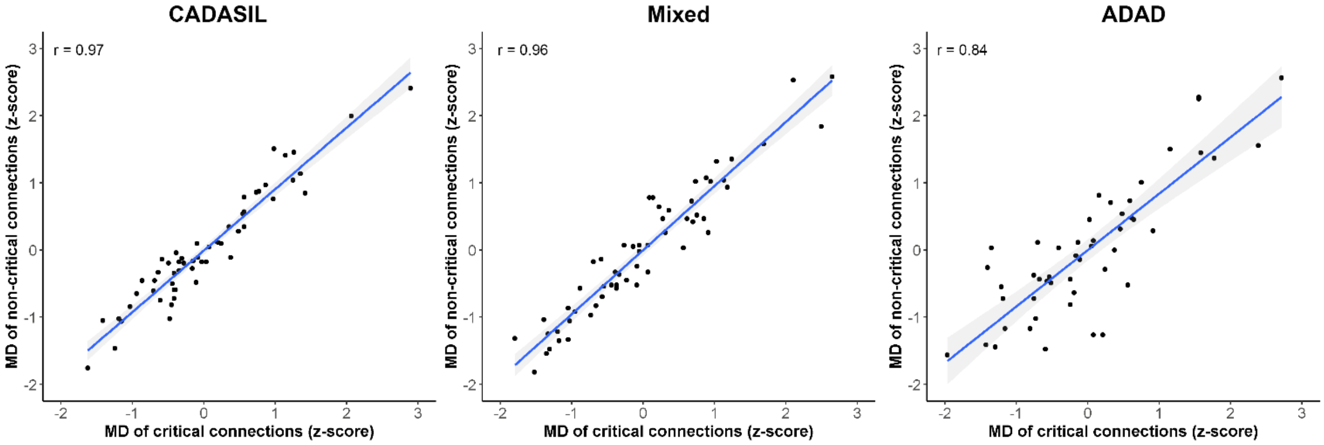

**Supplementary Figure 1.** Correlation coefficient between critical and non-critical connections

#### Supplementary Tables

##### Supplementary Table 1. Overview of study design

| | | Disease markers | | | | |
|---|---|---|---|---|---|---|
| | | **WMH** | **p-tau levels** | **Aβ status** | **Aβ levels** | **EYO** |
| **Network measures** | **MD-weighted** | | | | | |
| | Critical/non-critical connections | CADASIL/ Mixed | Mixed/ ADAD | Mixed/ ADAD | ADAD | ADAD |
| | Rich-club/ Feeder/Peripheral connections | CADASIL/ Mixed | Mixed/ ADAD | Mixed/ ADAD | ADAD | ADAD |
| | **FA-weighted** | | | | | |
| | Critical/non-critical connections | CADASIL/ Mixed | Mixed/ ADAD | Mixed/ ADAD | ADAD | ADAD |
| | Rich-club/ Feeder/Peripheral connections | CADASIL/ Mixed | Mixed/ ADAD | Mixed/ ADAD | ADAD | ADAD |

Aβ = amyloid beta; ADAD = Autosomal Dominant Alzheimer's Disease; EYO = Estimated years to symptom onset; FA= Fractional Anisotropy; MD = mean diffusivity; p-tau = phosphorylated tau; WMH = white matter hyperintensities.

**Supplementary Table 2. Acquisition parameters**

| | | CADASIL | Mixed | ADAD |
|---|---|---|---|---|
| **Scanner** | | Siemens Verio | Philips Achieva | Siemens systems |
| **T1** | TR [ms] | 2500 | 7.9 | 2300 |
| | TE [ms] | 437 | 4.5 | 2.95 |
| | Slice [mm] | 1 | 1 | 1.2 |
| | In-plane [mm] | 1 x 1 | 1 x 1 | 1.1 x 1.1 |
| **FLAIR** | TR [ms] | 5000 | 11000 | 9000 |
| | TE [ms] | 395 | 125 | 90 |
| | TI [ms] | 1800 | 2800 | 2500 |
| | Slice [mm] | 1 | 3 | 5 |
| | In-plane [mm] | 1 x 1 | 0.96 x 0.96 | 0.9 x 0.9 |
| **Diffusion** | TR [ms] | 12700 | 6600 | 6000 |
| | TE [ms] | 81 | 73 | 87 |
| | Slice [mm] | 2 | 2.5 | 2.5 |
| | In-plane [mm] | 2 x 2 | 1.72 x 1.72 | 2.5 x 2.5 |
| | b-value [s/mm$^2$] | 1000 | 1200 | 1000 |
| | directions | 30 | 45 | 64 |

ADAD = Autosomal dominant Alzheimer’s disease, FLAIR = Fluid attenuated inversion recovery, TE = Echo time, TR = Repetition time.

**Supplementary Tables 3a to 3e.**

The next five tables show the complete results of the robust linear regression models of the CADASIL, Mixed and Autosomal Dominant Alzheimer's Disease (ADAD) sample in relation to white matter hyperintensity volume (3a), Aβ positivity (3b), CSF Aβ42 (3c), CSF p-tau (3d) and estimated years from symptom onset (3e). For each of these disease burden markers we analyzed two different weightings for the connections (1) Mean diffusivity (MD) and (2) Fractional Anisotropy (FA) and two definitions of importance of connections (1) Critical and non-critical connections and (2) Rich club, feeder and peripheral connections. Results are presented with the standardized beta (St. B) with the 95% confidence interval (CI95) or the estimated marginal means (EMM) with the standard error (St. Error) and the corresponding *p*-value.

**Supplementary Table 3a. White matter hyperintensity volume**

| | **CADASIL** | | | **Mixed** | | |
|---|---|---|---|---|---|---|
| | St. B | CI95 | *p* | St. B | CI95 | *p* |
| **MD** | | | | | | |
| Critical | 0.70 | 0.53 – 0.88 | < 0.0001 | 0.71 | 0.54 – 0.88 | < 0.0001 |
| Non–critical | 0.70 | 0.53 – 0.87 | < 0.0001 | 0.79 | 0.64 – 0.93 | < 0.0001 |
| Rich club | 0.63 | 0.43 – 0.83 | < 0.0001 | 0.75 | 0.58 – 0.92 | < 0.0001 |
| Feeder | 0.67 | 0.49 – 0.84 | < 0.0001 | 0.75 | 0.61 – 0.89 | < 0.0001 |
| Peripheral | 0.73 | 0.56 – 0.89 | < 0.0001 | 0.79 | 0.63 – 0.94 | < 0.0001 |
| **FA** | | | | | | |
| Critical | -0.84 | -0.98 – -0.70 | < 0.0001 | -0.66 | -0.86 – -0.45 | < 0.0001 |
| Non-critical | -0.82 | -0.96 – -0.68 | < 0.0001 | -0.74 | -0.93 – -0.54 | < 0.0001 |
| Rich club | -0.71 | -0.89 – -0.52 | < 0.0001 | -0.76 | -0.94 – -0.59 | < 0.0001 |
| Feeder | -0.81 | -0.95 – -0.67 | < 0.0001 | -0.69 | -0.89 – -0.5 | < 0.0001 |
| Peripheral | -0.84 | -0.99 – -0.69 | < 0.0001 | -0.71 | -0.90 – -0.51 | < 0.0001 |

**Supplementary Table 3b. Aβ positivity**

| | **ADAD** | | | **Mixed** | | |
|---|---|---|---|---|---|---|
| | EMM | St. Error | *p* | EMM | St. Error | *p* |
| **MD** | | | | | | |
| Critical | -0.38 | 0.27 | 0.17 | -0.25 | 0.27 | 0.35 |
| Non-critical | -0.47 | 0.24 | 0.06 | -0.31 | 0.27 | 0.25 |
| Rich club | -0.37 | 0.24 | 0.15 | -0.38 | 0.27 | 0.16 |
| Feeder | -0.37 | 0.24 | 0.14 | -0.29 | 0.27 | 0.28 |
| Peripheral | -0.50 | 0.24 | 0.04 | -0.30 | 0.27 | 0.27 |
| **FA** | | | | | | |
| Critical | 0.63 | 0.32 | 0.05 | -0.02 | 0.27 | 0.94 |
| Non-critical | 0.96 | 0.30 | 0.003 | -0.03 | 0.27 | 0.91 |
| Rich club | 0.67 | 0.29 | 0.03 | 0.17 | 0.27 | 0.54 |
| Feeder | 0.91 | 0.28 | 0.002 | -0.05 | 0.27 | 0.86 |
| Peripheral | 0.89 | 0.30 | 0.006 | -0.02 | 0.27 | 0.95 |

**Supplementary Table 3c. CSF A$\beta^{42}$ levels**

| | ADAD | | |
|---|---|---|---|
| | St. B | CI95 | *p* |
| **MD** | | | |
| Critical | -0.25 | -0.48 – -0.02 | 0.04 |
| Non-critical | -0.31 | -0.54 – -0.08 | 0.01 |
| Rich club | -0.23 | -0.47 – 0.001 | 0.07 |
| Feeder | -0.26 | -0.51 – -0.01 | 0.04 |
| Peripheral | -0.32 | -0.55 – -0.1 | 0.008 |
| **FA** | | | |
| Critical | 0.34 | 0.04 – 0.65 | 0.03 |
| Non-critical | 0.45 | 0.15 – 0.76 | 0.006 |
| Rich club | 0.36 | 0.1 – 0.63 | 0.009 |
| Feeder | 0.46 | 0.2 – 0.73 | 0.001 |
| Peripheral | 0.41 | 0.11 – 0.72 | 0.01 |

**Supplementary Table 3d. Estimated years from symptom onset**

| | ADAD | | |
|---|---|---|---|
| | St. B | CI95 | *p* |
| **MD** | | | |
| Critical | 0.59 | 0.37 – 0.82 | < 0.0001 |
| Non-critical | 0.64 | 0.42 – 0.86 | < 0.0001 |
| Rich club | 0.47 | 0.24 – 0.71 | 0.00024 |
| Feeder | 0.63 | 0.40 – 0.87 | < 0.0001 |
| Peripheral | 0.64 | 0.41 – 0.87 | < 0.0001 |
| **FA** | | | |
| Critical | -0.07 | -0.36 – 0.20 | 0.58 |
| Non-critical | -0.21 | -0.51 – 0.09 | 0.17 |
| Rich club | -0.16 | -0.42 – 0.11 | 0.25 |
| Feeder | -0.34 | -0.62 – -0.06 | 0.02 |
| Peripheral | -0.12 | -0.41 – 0.18 | 0.43 |

**Supplementary Table 3e. CSF p-tau levels**

| | ADAD | | | Mixed | | |
|---|---|---|---|---|---|---|
| | St. B | CI95 | *p* | St. B | CI95 | *p* |
| **MD** | | | | | | |
| Critical | 0.33 | 0.13 – 0.54 | 0.003 | 0.07 | -0.27 – 0.41 | 0.7 |
| Non-critical | 0.21 | -0.03 – 0.45 | 0.09 | -0.07 | -0.40 – 0.26 | 0.66 |
| Rich club | 0.16 | -0.09 – 0.41 | 0.2 | 0.07 | -0.28 – 0.43 | 0.69 |
| Feeder | 0.22 | -0.02 – 0.46 | 0.08 | -0.03 | -0.35 – 0.29 | 0.85 |
| Peripheral | 0.23 | -0.01 – 0.46 | 0.05 | -0.06 | -0.44 – 0.33 | 0.77 |
| **FA** | | | | | | |
| Critical | 0.11 | -0.19 – 0.4 | 0.49 | 0.20 | -0.13 – 0.52 | 0.24 |
| Non-critical | -0.11 | -0.44 – 0.24 | 0.53 | 0.21 | -0.16 – 0.58 | 0.27 |
| Rich club | 0.12 | -0.15 – 0.40 | 0.38 | 0.03 | -0.33 – 0.40 | 0.85 |
| Feeder | -0.04 | -0.35 – 0.27 | 0.81 | 0.21 | -0.12 – 0.55 | 0.15 |
| Peripheral | -0.13 | 0.48 – 0.22 | 0.47 | 0.23 | -0.10 – 0.58 | 0.19 |

**Supplementary Tables 4a to 4e.**

The next five tables show the complete results of the robust linear regression models of the CADASIL, Mixed and Autosomal Dominant Alzheimer's Disease (ADAD) sample in relation to white matter hyperintensity volume (4a), Aβ positivity (4b), CSF Aβ42 (4c), CSF p-tau (4d) and estimated years from symptom onset (4e) corrected for age and sex. For each of these disease burden markers we analyzed two different weightings for the connections (1) Mean diffusivity (MD) and (2) Fractional Anisotropy (FA) and two definitions of importance of connections (1) Critical and non-critical connections and (2) Rich club, feeder and peripheral connections. Results are presented with the standardized beta (St. B) with the 95% confidence interval (CI95) or the estimated marginal means (EMM) with the standard error (St. Error) and the corresponding *p*-value.

**Supplementary Table 4a. White matter hyperintensity volume with age and sex correction**

| | **CADASIL** | | | **Mixed** | | |
|---|---|---|---|---|---|---|
| | St. B | CI95 | *p* | St. B | CI95 | *p* |
| **MD** | | | | | | |
| Critical | 0.68 | 0.46 – 0.92 | < 0.0001 | 0.71 | 0.50 – 0.93 | < 0.0001 |
| Non–critical | 0.64 | 0.42 – 0.87 | < 0.0001 | 0.79 | 0.64 – 0.93 | < 0.0001 |
| Rich club | 0.63 | 0.36 – 0.90 | < 0.0001 | 0.70 | 0.48 – 0.92 | < 0.0001 |
| Feeder | 0.63 | 0.39 – 0.86 | < 0.0001 | 0.74 | 0.55 – 0.93 | < 0.0001 |
| Peripheral | 0.68 | 0.46 – 0.91 | < 0.0001 | 0.78 | 0.59 – 0.98 | < 0.0001 |
| **FA** | | | | | | |
| Critical | -0.92 | -1.11 – -0.74 | < 0.0001 | -0.65 | -0.91 – -0.39 | < 0.0001 |
| Non-critical | -0.88 | -1.07 – -0.69 | < 0.0001 | -0.74 | -1.02 – -0.45 | < 0.0001 |
| Rich club | -0.73 | -0.98 – -0.48 | < 0.0001 | -0.71 | -0.95 – -0.48 | < 0.0001 |
| Feeder | -0.86 | -1.04 – -0.68 | < 0.0001 | -0.69 | -0.95 – -0.44 | < 0.0001 |
| Peripheral | -0.90 | -1.11 – -0.70 | < 0.0001 | -0.71 | -0.96 – -0.46 | < 0.0001 |

**Supplementary Table 4b. Aβ positivity with age and sex correction**

| | ADAD | | | Mixed | | |
|---|---|---|---|---|---|---|
| | EMM | St. Error | *p* | EMM | St. Error | *p* |
| **MD** | | | | | | |
| Critical | -0.11 | 0.28 | 0.69 | -0.21 | 0.24 | 0.39 |
| Non-critical | -0.16 | 0.23 | 0.49 | -0.23 | 0.24 | 0.34 |
| Rich club | -0.15 | 0.26 | 0.56 | -0.31 | 0.23 | 0.19 |
| Feeder | -0.07 | 0.26 | 0.56 | -0.23 | 0.24 | 0.33 |
| Peripheral | -0.20 | 0.23 | 0.40 | -0.22 | 0.24 | 0.36 |
| **FA** | | | | | | |
| Critical | 0.72 | 0.35 | 0.05 | -0.04 | 0.26 | 0.88 |
| Non-critical | 0.87 | 0.33 | 0.01 | -0.09 | 0.26 | 0.73 |
| Rich club | 0.72 | 0.32 | 0.03 | 0.10 | 0.25 | 0.67 |
| Feeder | 0.84 | 0.31 | 0.01 | -0.09 | 0.26 | 0.73 |
| Peripheral | 0.83 | 0.34 | 0.02 | -0.07 | 0.26 | 0.79 |

**Supplementary Table 4c. CSF Aβ$^{42}$ levels with age and sex correction**

| | ADAD | | |
|---|---|---|---|
| | St. B | CI95 | *p* |
| **MD** | | | |
| Critical | -0.11 | -0.40 – 0.18 | 0.46 |
| Non-critical | -0.16 | -0.40 – 0.08 | 0.20 |
| Rich club | -0.13 | -0.39 – 0.13 | 0.35 |
| Feeder | -0.09 | -0.33 – 0.15 | 0.47 |
| Peripheral | -0.18 | -0.42 – -0.05 | 0.14 |
| **FA** | | | |
| Critical | 0.36 | 0.04 – 0.68 | 0.03 |
| Non-critical | 0.42 | 0.08 – 0.76 | 0.02 |
| Rich club | 0.38 | 0.01 – 0.68 | 0.01 |
| Feeder | 0.43 | 0.14 – 0.74 | 0.007 |
| Peripheral | 0.38 | 0.04 – 0.72 | 0.03 |

**Supplementary Table 4d. Estimated years from symptom onset with age and sex correction**

| | ADAD | | |
|---|---|---|---|
| | St. B | CI95 | *p* |
| **MD** | | | |
| Critical | 0.60 | 0.20 – 1.00 | 0.005 |
| Non-critical | 0.48 | 0.10 – 0.85 | 0.016 |
| Rich club | 0.34 | -0.06 – 0.76 | 0.10 |
| Feeder | 0.51 | 0.12 – 0.89 | 0.013 |
| Peripheral | 0.47 | 0.08 – 0.86 | 0.02 |
| **FA** | | | |
| Critical | -0.17 | -0.66 – 0.31 | 0.49 |
| Non-critical | 0.08 | -0.44 – 0.60 | 0.76 |
| Rich club | 0.001 | -0.48 – 0.49 | 0.99 |
| Feeder | -0.06 | -0.59 – 0.47 | 0.81 |
| Peripheral | 0.12 | -0.40 – 0.65 | 0.65 |

**Supplementary Table 4e. CSF p-tau levels with age and sex correction**

| | ADAD | | | Mixed | | |
|---|---|---|---|---|---|---|
| | St. B | CI95 | *p* | St. B | CI95 | *p* |
| **MD** | | | | | | |
| Critical | 0.23 | -0.02 – 0.48 | 0.09 | 0.01 | -0.29 – 0.32 | 0.92 |
| Non-critical | -0.01 | -0.27 – 0.25 | 0.93 | -0.13 | -0.42 – 0.15 | 0.38 |
| Rich club | 0.05 | -0.24 – 0.35 | 0.72 | 0.004 | -0.29 – 0.29 | 0.98 |
| Feeder | -0.0003 | -0.25 – 0.25 | 0.99 | -0.05 | -0.33 – 0.23 | 0.74 |
| Peripheral | 0.03 | -0.22 – 0.28 | 0.82 | -0.11 | -0.41 – 0.19 | 0.47 |
| **FA** | | | | | | |
| Critical | 0.21 | -0.12 – 0.54 | 0.22 | 0.22 | -0.07 – 0.51 | 0.16 |
| Non-critical | 0.01 | -0.37 – 0.40 | 0.96 | 0.23 | -0.06 – 0.53 | 0.13 |
| Rich club | 0.20 | -0.13 – 0.54 | 0.24 | 0.09 | -0.19 – 0.36 | 0.53 |
| Feeder | 0.14 | -0.21 – 0.50 | 0.43 | 0.22 | -0.06 – 0.51 | 0.13 |
| Peripheral | -0.025 | -0.43 – 0.38 | 0.90 | 0.23 | -0.07 – 0.53 | 0.15 |

**Supplementary Table 5. Sensitivity analysis for ADAD sample with scanner correction**

| | Estimated years of onset | | | CSF Aβ levels | | |
|---|---|---|---|---|---|---|
| | St. B | CI95 | *p* | St. B | CI95 | *p* |
| **MD** | | | | | | |
| Critical | 0.65 | 0.42 – 0.89 | <0.001 | -0.26 | -0.50 – -0.01 | 0.04 |
| Non-critical | 0.69 | -0.27 – 0.25 | <0.001 | -0.32 | -0.56 – -0.08 | 0.01 |
| Rich club | 0.53 | 0.29 – 0.76 | <0.001 | -0.26 | -0.50 – -0.03 | 0.03 |
| Feeder | 0.70 | 0.45 – 0.94 | <0.001 | -0.28 | -0.52 – -0.03 | 0.04 |
| Peripheral | 0.69 | 0.45 – 0.93 | <0.001 | -0.32 | -0.54 – -0.10 | 0.007 |

**Supplementary Table 6. DIAN consortium**

| Last Name | First | Affiliation |
|---|---|---|
| Allegri | Ricardo | FLENI Institute of Neurological Research (Fundacion para la Lucha contra las Enfermedades Neurologicas de la Infancia) |
| Bateman | Randy | Washington University in St. Louis School of Medicine |
| Bechara | Jacob | Neuroscience Research Australia |
| Benzinger | Tammie | Washington University in St. Louis School of Medicine |
| Berman | Sarah | University of Pittsburgh |
| Bodge | Courtney | Brown University-Butler Hospital |
| Brandon | Susan | Washington University in St. Louis School of Medicine |
| Brooks | William (Bill) | Neuroscience Research Australia |
| Buck | Jill | Indiana University |
| Buckles | Virginia | Washington University in St. Louis School of Medicine |
| Chea | Sochenda | Mayo Clinic Jacksonville |
| Chhatwal | Jasmeer | Brigham and Women's Hospital–Massachusetts General Hospital |
| Chrem | Patricio | FLENI Institute of Neurological Research (Fundacion para la Lucha contra las Enfermedades Neurologicas de la Infancia) |
| Chui | Helena | University of Southern California |
| Cinco | Jake | University College London |
| Clifford | Jack | Mayo Clinic Jacksonville |
| Cruchaga | Carlos | Washington University in St. Louis School of Medicine |
| Donahue | Tamara | Washington University in St. Louis School of Medicine |
| Douglas | Jane | University College London |
| Edigo | Noelia | FLENI Institute of Neurological Research (Fundacion para la Lucha contra las Enfermedades Neurologicas de la Infancia) |

| | | |
|---|---|---|
| Erekin-Taner | Nilufer | Mayo Clinic Jacksonville |
| Fagan | Anne | Washington University in St. Louis School of Medicine |
| Farlow | Marty | Indiana University |
| Fitzpatrick | Colleen | Brigham and Women's Hospital-Massachusetts |
| Flynn | Gigi | Washington University in St. Louis School of Medicine |
| Fox | Nick | University College London |
| Franklin | Erin | Washington University in St. Louis School of Medicine |
| Fujii | Hisako | Osaka City University |
| Gant | Cortaiga | Washington University in St. Louis School of Medicine |
| Gardener | Samantha | Edith Cowan University, Perth |
| Ghetti | Bernardino | Indiana University |
| Goate | Alison | Icahn School of Medicine at Mount Sinai |
| Goldman | Jill | Columbia University |
| Gordon | Brian | Washington University in St. Louis School of Medicine |
| Graff-Radford | Neill | Mayo Clinic Jacksonville |
| Gray | Julia | Washington University in St. Louis School of Medicine |
| Groves | Alexander | Washington University in St. Louis School of Medicine |
| Hassenstab | Jason | Washington University in St. Louis School of Medicine |
| Hoechst-Swisher | Laura | Washington University in St. Louis School of Medicine |
| Holtzman | David | Washington University in St. Louis School of Medicine |
| Hornbeck | Russ | Washington University in St. Louis School of Medicine |
| Houeland DiBari | Siri | German Center for Neurodegenerative Diseases (DZNE) Munich |
| Ikeuchi | Takeshi | Niigata University |

| | | |
|---|---|---|
| Ikonomovic | Snezana | University of Pittsburgh |
| Jerome | Gina | Washington University in St. Louis School of Medicine |
| Jucker | Mathias | German Center for Neurodegenerative Diseases (DZNE) Tubingen |
| Karch | Celeste | Washington University in St. Louis School of Medicine |
| Kasuga | Kensaku | Niigata University |
| Kawarabayashi | Takeshi | Hirosaki University |
| Klunk | William (Bill) | University of Pittsburgh |
| Koeppe | Robert | University of Michigan |
| Kuder-Buletta | Elke | German Center for Neurodegenerative Diseases (DZNE) Tubingen |
| Laske | Christoph | German Center for Neurodegenerative Diseases (DZNE) Tubingen |
| Lee | Jae-Hong | Asan Medical Center |
| Levin | Johannes | German Center for Neurodegenerative Diseases (DZNE) Munich |
| Martins | Ralph | Edith Cowan University |
| Mason | Neal Scott | University of Pittsburgh Medical Center |
| Masters | Colin | University of Melbourne |
| Maue-Dreyfus | Denise | Washington University in St. Louis School of Medicine |
| McDade | Eric | Washington University in St. Louis School of Medicine |
| Mori | Hiroshi | Osaka City University |
| Morris | John | Washington University in St. Louis School of Medicine |
| Nagamatsu | Akem | Tokyo University |
| Neimeyer | Katie | Columbia University |
| Noble | James | Columbia University |
| Norton | Joanne | Washington University in St. Louis School of Medicine |

| | | |
|---|---|---|
| Perrin | Richard | Washington University in St. Louis School of Medicine |
| Raichle | Marc | Washington University in St. Louis School of Medicine |
| Renton | Alan | Icahn School of Medicine at Mount Sinai |
| Ringman | John | University of Southern California |
| Roh | Jee Hoon | Asan Medical Center |
| Salloway | Stephen | Brown University-Butler Hospital |
| Schofield | Peter | Neuroscience Research Australia |
| Shimada | Hiroyuki | Osaka City University |
| Sigurdson | Wendy | Washington University in St. Louis School of Medicine |
| Sohrabi | Hamid | Edith Cowan University |
| Sparks | Paige | Brigham and Women's Hospital-Massachusetts |
| Suzuki | Kazushi | Tokyo University |
| Taddei | Kevin | Edith Cowan University |
| Wang | Peter | Washington University in St. Louis School of Medicine |
| Xiong | Chengjie | Washington University in St. Louis School of Medicine |
| Xu | Xiong | Washington University in St. Louis School of Medicine |
